# Estimated incidence and number of new cases with prenatal alcohol exposure and fetal alcohol spectrum disorders in Finland 1990-2025

**DOI:** 10.64898/2026.05.09.26352787

**Authors:** Mirjami Jolma, Mikko Koivu-Jolma, Mika Gissler, Anne Sarajuuri, Ilona Autti-Rämö

**Affiliations:** Faculty of Medicine, The University of Helsinki, Helsinki, Finland; Division of Child Neurology, Päijät-Häme Central Hospital, Lahti, Finland; Faculty of Science, The University of Helsinki, Helsinki, Finland; Department of Environmental and Biological Sciences, The University of Eastern Finland, Joensuu, Finland; Department of Data and Analytics, THL Finnish Institute for Health and Welfare, Helsinki, Finland; Department of Molecular Medicine and Surgery, Karolinska Institute and Academic Primary Health Care Centre, Region Stockholm, Stockholm, Sweden; Research Centre for Child Psychiatry, University of Turku, Turku, Finland; Division of Child Neurology, University of Helsinki, New Children’s hospital, Helsinki, Finland

## Abstract

**Background:** Fetal alcohol spectrum disorders (FASD) arising from prenatal alcohol exposure (PAE) are the leading preventable cause of neurobehavioral disorders. Early pregnancy is particularly vulnerable to ethanol toxicity, yet alcohol use often continues until pregnancy recognition. In Finland, national incidence estimates of PAE and FASD remain limited.

**Objective:** To estimate the incidence of any PAE and heavy PAE in Finland between 1990 and 2025, and to model the annual number of children with FASD born in or immigrating to Finland.

**Methods:** We developed a mathematical modelling framework integrating studies on alcohol use during pregnancy in Finland, biomarker-based estimates of heavy PAE, national population statistics, and international active case ascertainment studies on FASD prevalence. Incidence of any PAE was estimated from self-reported alcohol use, including pre-recognition exposure. Heavy PAE was estimated by combining binge-drinking prevalence, delayed pregnancy recognition, biomarker data and anonymous self-reports. FASD incidence was modelled using two approaches: 1) an international multiplier linking FASD prevalence to heavy episodic drinking prevalence among women, and 2) a conventional epidemiological ratio between any PAE and FASD. Immigration and international adoption were incorporated.

**Results:** Self-reported alcohol use during pregnancy declined following abstinence recommendations in the early 2000s, while pre-recognition use remained relatively stable. Heavy PAE decreased from 9% (uncertainty range 7-11%) in the 1990s to 6% (4-8%) in the early 2020s. Any PAE declined from 75% (60-85%) to 32% (26-38%). Modelled FASD incidence showed similar decreasing trends, ranging from 6.8% to 5.6% (multiplier model), and from 6% to 3% (any PAE-based model).

**Conclusion:** PAE remains common in Finland, and the burden of FASD is substantial despite declining trends. Additional biomarker-based studies of PAE and active case ascertainment of FASD are needed to refine current estimates. Strengthened public health efforts to reduce PAE, including the efforts before recognition of pregnancy, are essential.

## Introduction

Fetal alcohol spectrum disorders (FASD) are the leading preventable cause of intellectual disability (O’Leary et al. 2013), developmental and neurobehavioral disorders, congenital anomalies and other lifelong problems caused by prenatal alcohol exposure (PAE) [1]. No safe timing or amount of alcohol use during pregnancy has been established [1,2]. Ethanol readily crosses biological membranes [3] and even low levels of early PAE can cause discernible permanent alterations in brain and facial development [4]. Experimental evidence indicates that the earliest stages of pregnancy are the most vulnerable [5] and ethanol can affect embryonic development and placental formation even before implantation, causing changes in gene regulation and epigenome [6,7]. The facial dysmorphism characteristic of fetal alcohol syndrome arises from PAE during gastrulation [8], corresponding to approximately the fifth gestational week, when many pregnancies remain still unrecognized [9]. The developing brain is vulnerable to the toxic effects of ethanol during the whole pregnancy [1,5,10].

The risk of FASD increases with higher levels of PAE, particularly heavy episodic drinking or binge drinking, which produces high peak ethanol concentrations in the embryo or fetus [1,5]. Although pregnancy is often associated with reduced regular alcohol use, binge drinking may persist among some users [11]. In Finland, alcohol use has traditionally been characterized by heavy episodic drinking, and alcohol-related morbidity among women has exceeded European averages since mid-1990s [12]. A global review and meta-analysis including studies from 1984 to 2014 relying on self-reporting and assuming a constant rate of PAE over time, estimated that 25.2% of European pregnancies involved any degree of PAE and 2.7% involved binge drinking. [13]. Using these estimates and then-available prevalence studies, diagnosable FASD prevalence was estimated at constant rate of 1/13 of any PAE globally [14]. However, this approach assumes homogenous risk across exposure patterns, ignores timing and dose effects and yields implausible estimates in settings with high observed FASD prevalence.

The estimates of PAE and FASD and the rate between them depend not only on differences in alcohol use cultures but also on data collection methods, prevalence study protocols and applied FASD diagnostic criteria, which differ substantially in sensitivity [15]. Reliability of data on PAE is bound to depend on attitudes towards alcohol use during pregnancy and methods of data collection. Self-reported PAE is strongly influenced by stigma following recommendations for complete abstinence during pregnancy. Biomarker studies indicate that only a very small proportion of women with evidence of PAE disclose alcohol use [16], unless anonymity is ensured [17]. European biomarker studies demonstrate substantially higher PAE prevalence rates than the prior estimate for Europe with varying drinking patterns: e.g. in Spain maternal hair ethyl glucuronide (EtG) indicated any detectable PAE in 63%, but heavy PAE only in 2% [16]. In Poland maternal hair EtG indicated any PAE in 50% but heavy PAE in 10%, not depending on socioeconomic status [18]. These factors complicate comparisons across countries and time periods and contribute to uncertainty in prevalence estimates.

In Finland, the prevalence of PAE and FASD remain poorly characterized. The only crude Finnish national estimate from the late 1980s by an expert in the field (Ilona Autti-Rämö) suggests that 600-3000 children with FASD were born annually [19,20]. However, only 556 cases of dysmorphic FASD were recorded in the national register of congenital malformations between 1990 and 2017 (Register for congenital malformations), even though alcohol use among women has increased. This indicates extensive under-recognition [21], consistent with findings in other countries [22,23]. Although most individuals with FASD remain undiagnosed, they likely receive multiple other diagnoses for neurodevelopmental, psychiatric and somatic conditions associated with FASD [24,25] even while etiology remains unrecognized.

The current gold-standard for estimating FASD prevalence is in-school active case ascertainment protocol [26] based on earlier active case ascertainment studies [27,28]. This approach has yielded higher prevalence estimates than earlier passive surveillance [26] suggesting that earlier estimates may have underestimated the population burden of FASD in Europe. The in-school active case ascertainment approach is resource-intensive, limited to specific age groups and still subject to participation, screening and exclusion biases. It also cannot capture temporal trends without repeated studies. In-school studies from countries with women’s alcohol use patterns broadly comparable to Finland, have reported observed FASD prevalence among participants ranging from 1.8% to 12.2%, depending on area, applied diagnostic criteria, participation rates and study period [23,29–35].

Given the absence of national active case ascertainment studies and the need to estimate incidence across multiple birth cohorts, alternative approaches are required. Mathematical modelling that integrates heterogenous data sources on PAE, with the emphasis on biomarkers and anonymous self-reporting and indicators of heavy alcohol use, together with prevalence estimates from comparable in-school studies, provides a pragmatic method for estimating the population burden of PAE and FASD over time. Such approach allows explicit handling of uncertainty and estimation of trends that cannot be captured by point-prevalence studies alone.

The aims of this study were:

1. To estimate, using available data and mathematical modelling, the incidence and number of individuals born with any PAE and with heavy PAE in Finland between 1990 and 2025.
2. To estimate the annual increase of children with FASD in Finland during this period.

## Methods and data

### Definitions and abbreviations

FASD= fetal alcohol spectrum disorders, PAE= prenatal alcohol exposure, AUD = alcohol use disorder, BrP = before recognition of pregnancy, ArP = after recognition of pregnancy.

### Conceptual framework and assumptions

PAE was assumed to arise through three non-mutually exclusive mechanisms:

1. Alcohol use before recognition of pregnancy
2. Continued alcohol use during pregnancy due to maternal AUD
3. Continued alcohol use due to low self-perceived risk of fetal harm

Following the introduction of national recommendations for complete abstinence during pregnancy (explicit since 2004 in Finland), non-anonymous self-reported PAE ArP was assumed unreliable. Accordingly, particularly post-2004 estimates prioritize biomarkers and fully anonymous self-reporting.

The framework reflects the assumptions that, at the population level, PAE prevalence is associated with alcohol use prevalence, drinking patterns and prevalence of delayed pregnancy awareness among women. FASD depends on PAE, with highest risk associated with heavy early exposure, particularly heavy episodic drinking. Thus, incidences of any PAE and heavy PAE, including BrP, were estimated separately. With decreasing birth rates and increasing immigration, migration was incorporated to account for children with FASD born abroad who later immigrated, either through international adoption or with their birth parents.

### Data sources

Parameters used as input for modelling (Table 1) were adapted from:

1. Published studies on PAE in Finland, in English and Finnish
2. National population statistics
3. Annual estimates of any alcohol use and heavy episodic drinking among women (WHO)
4. European and North American in-school active case ascertainment studies on FASD prevalence

**Table 1.**
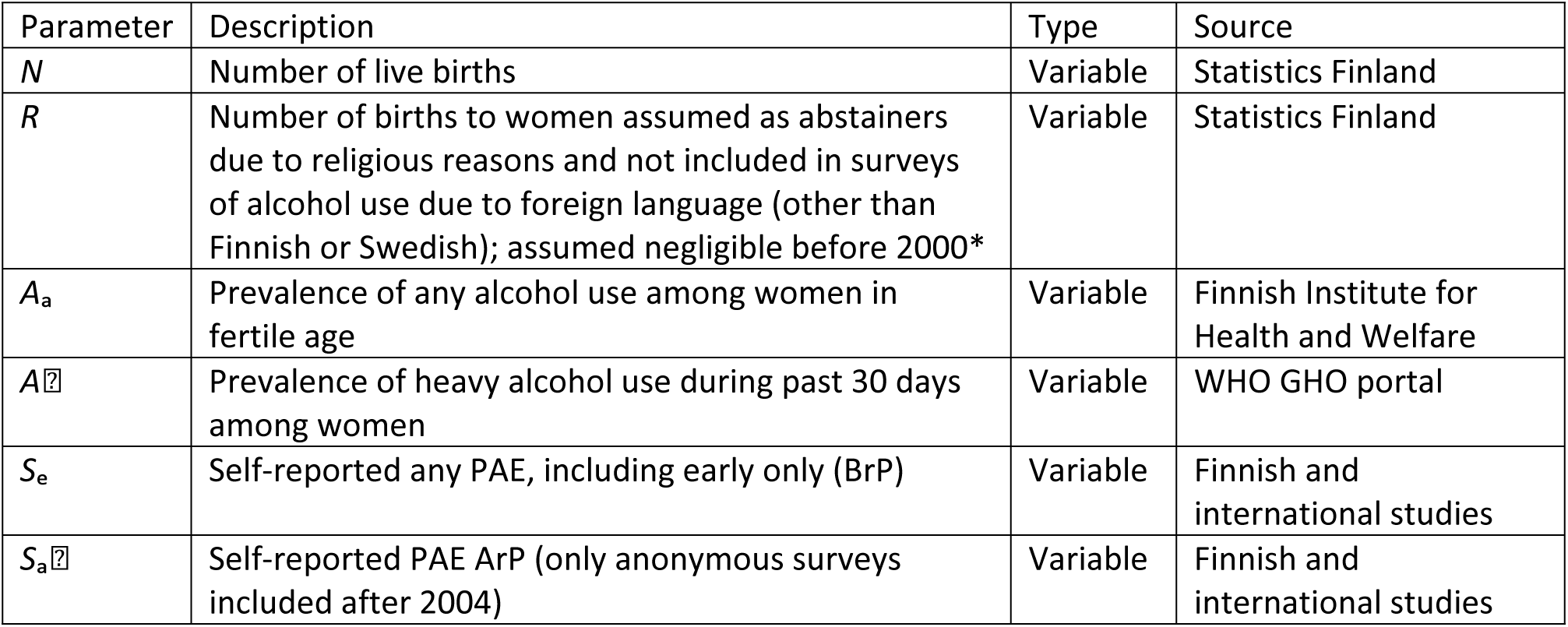
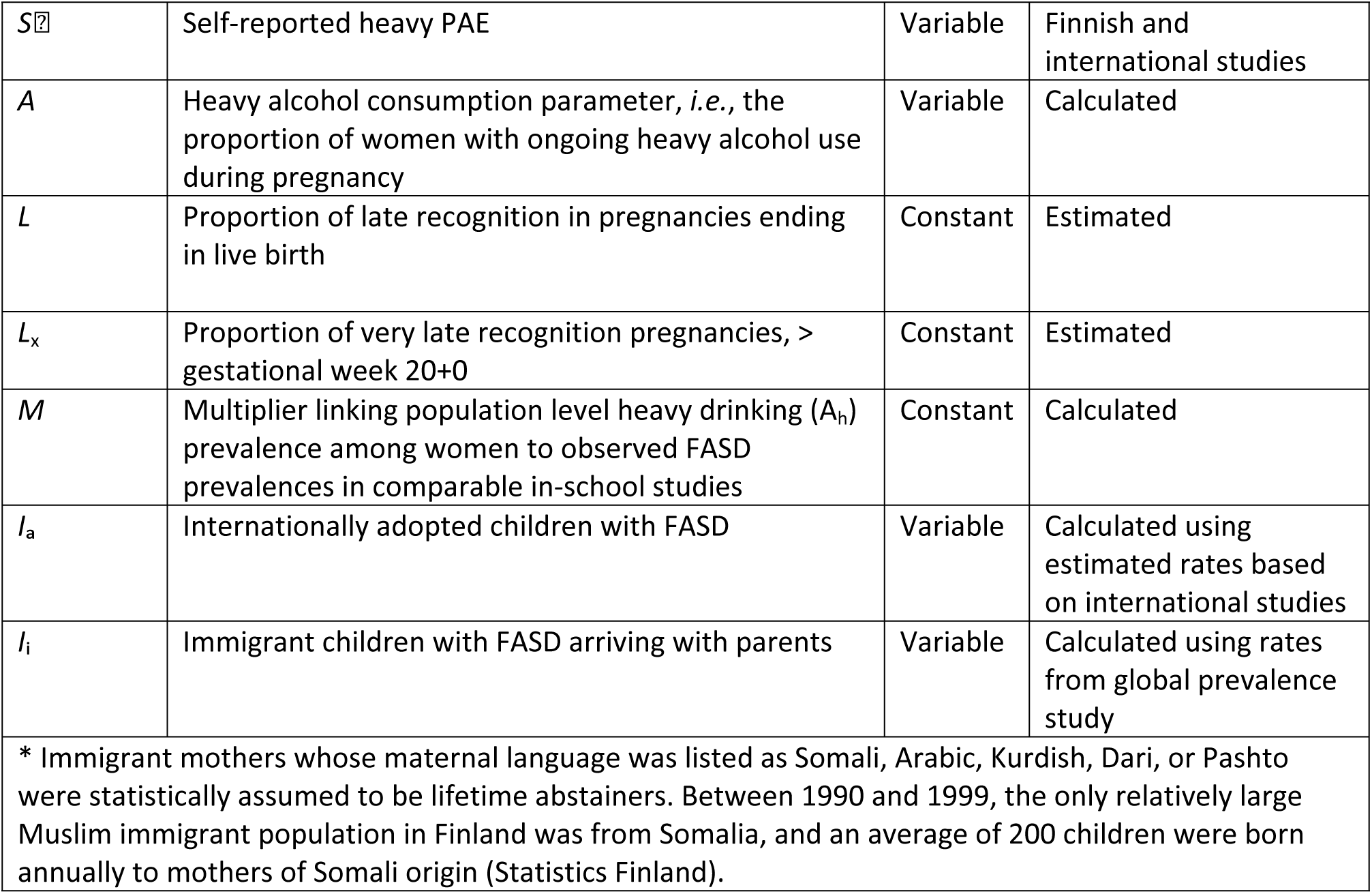
Parameters.

Due to a very small number of biomarker studies and sporadic fully anonymous self-reporting studies, we could not use the more reliable, albeit not very sensitive, biomarkers and anonymous self-reporting but had to combine data from heterogenous sources. When multiple sources provided divergent estimates, greater weight was given to biomarker-based and anonymous self-reporting data. From the available data, we extracted trends using linear modelling.

### Included studies for data collection

For PAE studies, we searched in Google Scholar and PubMed, both in English and in Finnish, for studies on alcohol use in pregnancy (English search terms: Finland, alcohol, pregnancy, PAE, prenatal alcohol exposure, prevalence, incidence; Finnish search terms: alkoholi [alcohol], raskaus [pregnancy], äitiysneuvola [prenatal clinic], päihteet [substance use]). For each included study we also searched for references indicating other studies on the subject. The Finnish studies with very small population sizes (<100) conducted by students at universities of applied sciences were excluded. Included studies are described in Table 2 and listed according to their type, either self-reporting or biomarker, and in self-reporting studies the degree of anonymity in data collection is listed.

**Table 2.** PAE prevalence studies in Finland.

| Study and type | Anonymity and method of information gathering | Number of participants/ recruited (participation rate if available) | Period of data collection | Alcohol use during pregnancy and timing<br>BrP =before recognition of pregnancy<br>ArP = after recognition of pregnancy |
| --- | --- | --- | --- | --- |
| Self-reporting studies |  |  |  |  |
| Mårdby et al. 2017 [36] | Anonymous reporting | 572 | 2011-2012 | 79/572 (14%) ArP |
| Komulainen et al. 2017 [37] | Anonymous reporting | 380/763 (49.8%) | 2012 | 207/380 (54.5%) any<br>81/380 (21.3%) 3+ drinks at a time<br>16/380 (4.2%) 7+ drinks at a time |
| Voutilainen et al. 2022 [38] | Semi-anonymous reporting | 15461/21472 (72%) | 2009-2018 | 26% BrP<br>4.5% ArP |
| Lehtinen and Ekblad 2023 [39] | Non-anonymous reporting | 248/589 (42.1%) | 2016-2019 | 55% BrP<br>0% ArP |
| Raatikainen et al. 2007 [40] | Non-anonymous questionnaire and interview, plus a database | 25591 | 1989-2001 | Pregnancy week 20<br>47.6% BrP of smokers<br>6.9% ArP of smokers<br>35.2% BrP of non-smokers<br>2.9% ArP of non-smokers<br>39.4% all BrP<br>3.5% all ArP |
| Kurki et al. 2000 [41] | Non-anonymous Reporting | 623 | NA | Pregnancy weeks 10-17<br>Any use 350/623 (56.2%)<br>at least weekly use 56/623 (9.0%) |
| Halmesmäki et al. 1987 [42] | Information not available | 530 | NA | 90% any use<br>10% more than 2 drinks/week |
| Halmesmäki and Kinnunen 1993 [43] | Anonymous questionnaire | 788/3364 (23.4%) | 1990 | 35.8% any use ArP<br>7% monthly use<br>2% 3 or more drinks at a time |
| Finnish Institute for Health and Welfare [44] | Semi-anonymous questionnaire | 8977/17964 (50%) | 2020 | 1.9% ArP |
| Raitasalo et al. 2015 [45] | National registers<br>Alcohol use disorder (AUD) diagnoses | 54519 (all mothers that gave birth in 2002) | Children born 2002, maternal AUD diagnoses 1998–2009 | 833/54519 (1.5%) of pregnancies in mothers with AUD diagnosis |
| Niemelä et al. 2016 [46] | Non-anonymous questionnaire and maternal serum biobank samples | 806 (50% with gestational diabetes, part of FinGeDi-study) | 2009-2012 | 95/806 (11.8%) any use self-reported. As a group, also those not reporting any alcohol use had significantly higher GGT and GGT-CDT levels compared to pregnant lifelong abstainers |
| Biomarker studies |  |  |  |  |
| Häkkinen et al. 2024 S-Peth [47] | Anonymous blood samples | 3000 | 2023 | Pregnancy weeks 8-12<br>PEth > 0 ng/mL 253/3000 (8.4%)<br>PEth ≥ 2 ng/mL in 156/3000 (5.2%)* |
| Jolma et al. 2023 [48] | Anonymous urine samples (short detection window) | 505 | 2020–2021 | EtG > 100 ng/mL **<br>12/68 (17.6%) of HAL clinic samples<br>31/437 (7.1%) of all other samples<br>EtG > 300ng/ml<br>4/212 (1.9%) general prenatal clinic |
| *PEth elevates only due to alcohol in blood, low amounts of alcohol typically do not cause detectable PEth levels (Kwak et. 2014), 2ng/ml can be considered significant during pregnancy [47]. ** During pregnancy urine EtG cut-off of 100ng/ml has been recommended to detect PAE [49,50]. |  |  |  |  |

For FASD prevalence estimation, first we searched Google Scholar and PubMed and references of epidemiologic FASD studies (using search terms: FASD prevalence, active case ascertainment, in-school) and collected all found in-school active case ascertainment studies from Europe and North America.

Included studies are listed in Table 3. Applied diagnostic criteria are listed and the emphasis is on studies using IOM 2005 and IOM 2016 diagnostic guidelines, which are bolded, as those are used in Finland. We collected the years when children participating in each study were born and the corresponding women’s alcohol use indicator (prevalence of women with heavy episodic alcohol use during past 30 days = *A*_h_) at one year before the birth years with the estimated FASD prevalence.

**Table 3.**
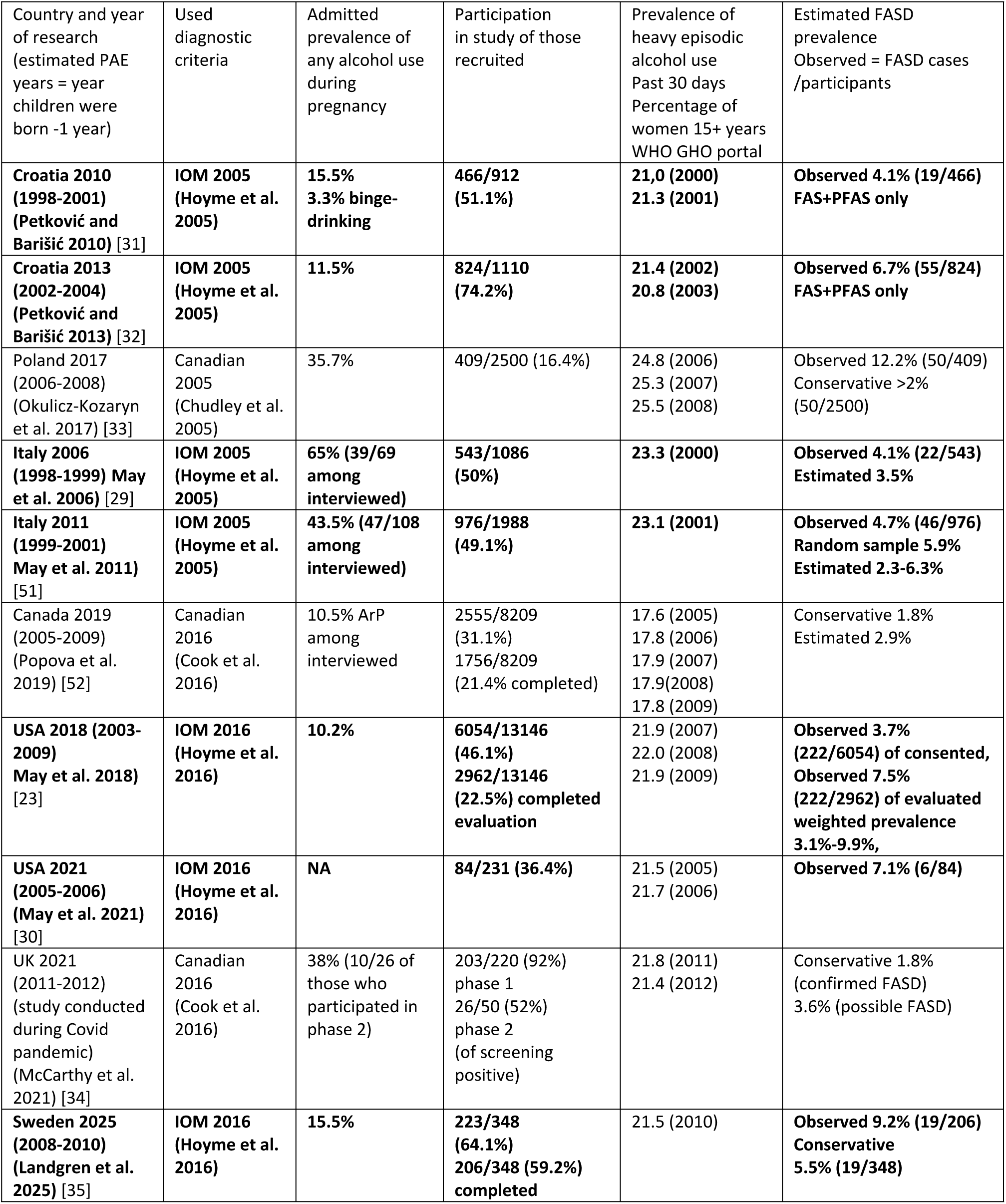
Prevalence of FASD in countries comparable to Finland and women’s heavy episodic alcohol use indicator (A_h_ in models) for years of gestation of study participants. Studies using IOM 2016 diagnostic guidelines are bolded.

### Estimation of PAE incidence

Yearly incidence of any PAE (*P*_a_(*t*)) was estimated using self-reported PAE including BrP:

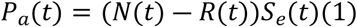

where *N*(*t*) stands for the number of live births and *R*(*t*) for the estimated number non-Finnish/Swedish speaking abstainers giving birth during a single year. *S*_e_(*t*) is the proportion of pregnant women who reported alcohol use at the same year.

We estimated yearly incidence of heavy PAE using a model combining biomarker-based data with reported alcohol use. Due to the unreliability of self-reporting, we assigned double weight for the biomarker-based data and anonymous self-reports compared to non-anonymous self-reports.

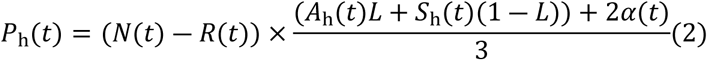

Here *α* is the proportion of the expecting mothers with ongoing heavy alcohol use during pregnancy (see section Estimation of variables), *A*_h_ is the prevalence of binge drinking during past 30 days among women, *L* is the proportion of late awareness of pregnancy and *S*_h_ is the self-reported heavy alcohol use during pregnancy. The resulting estimate is conservative as the biomarker detection window does not extend to very early pregnancy and due to substantially under-reported heavy use during pregnancy.

### Estimation of FASD incidence

Estimating FASD differed from simple estimation of PAE or even heavy PAE, as no in-school active case ascertainment prevalence estimation studies have been conducted for Finland. Consequently, we had to base our estimation on observed FASD prevalences in available European and North American in-school studies (in Sweden, Poland, Croatia, Italy, UK, US and Canada).

We compared two simple models for FASD incidence estimation for each year. The first, which here we call the M model, was based on the ratio of binge drinking among women and FASD prevalence according to international studies.

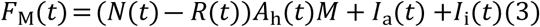

In this model *F*_M_ stands for yearly FASD prevalence. As in the PAE estimations, *N* stands for the number of live births, *R* for the estimated number of non-Finnish/Swedish speaking abstainers not included in statistics and *A*_h_ for the prevalence of heavy alcohol use during past 30 days among women. *M* is a country multiplier, which is described in the section Estimation of constants. In addition, *I*_a_ and *I*_i_ are the estimated numbers of internationally adopted children with FASD and immigrant children with FASD arriving with parents, respectively. The estimation processes for *I*_a_ and *I*_i_ are described in the section Estimation of immigrant children with FASD. All variables change annually, whereas *M* is constant.

The second model was based on the prior epidemiologic ratio by Lange et al. [14] and thus we call it the L model here.

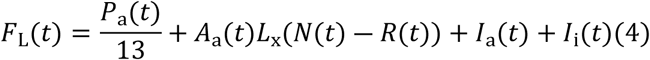

In L model *F*_L_ stands for FASD prevalence within the age cohort and *P*_a_ (Eq. 1) for incidence of any PAE within the cohort. In addition to the simple ratio, *A*_a_ is the prevalence of any alcohol use among women in childbearing age and *L*_x_ is the proportion of very late (after gestational week 20) recognized pregnancies not included in studies on self-reporting PAE conducted during earlier gestation. Variables *N*, *R*, *I*_a_ and *I*_i_ are the same as in equation (1). All variables change annually, whereas *L*_x_ is constant.

### Estimation of variables

Due to the sparse data, we had to extrapolate and make assumptions about the prevalence of alcohol use. Data on general alcohol use prevalence by women aged 20 – 34 and 35 – 44 years (*A*_a_) was available for selected years between 1990 and 2023[53], whereas yearly data on prevalence of binge drinking (*A*_h_), by WHO GHO (Global Health Observatory) as estimates comparable to other countries, was available for years 2000 – 2019 [54]. We defined *A*_a_ as the mean prevalence of the two age groups (20 – 34 and 35 – 44 years). We set the missing 1990 *A*_h_ to the mean of known *A*_h_ values. Because the biomarker studies presented two different prevalence numbers for alcohol use depending on the used cut-off, we used their mean value. The proportion of the expecting mothers with ongoing heavy alcohol use during pregnancy (*α*) was formed by combining self-reported heavy use data before year 2004 with biomarker data after year 2004. Following this, we used first order linear models to predict the data points for years 1987 – 2024 to be used in final PAE and FASD modelling efforts. Supplementary files to reproduce the adjustments and the results include data files (S1 Dataset) and the R source file (S2 Source).

### Estimation for constants

#### Prevalence of late pregnancy awareness

As PAE during gastrulation appears particularly harmful to developing embryo [6–8], pregnancy awareness and cessation of alcohol use after gestational week 4+0 can be considered “late”. Modern pregnancy tests can detect pregnancy earliest at gestational age 3+3 weeks. Traditionally, awareness before missed menstruation, typically at gestational week 4+0, was not possible, placing all women continuing alcohol use until awareness at risk for early PAE, particularly those with irregular periods.

Unintended pregnancy is a risk factor for early PAE, increasing risk for both continued alcohol use until recognition and delayed awareness. Unintended pregnancies include unplanned, mistimed and ambivalent pregnancies or belief of low fertility. Unintended pregnancies ending in abortion are excluded as they cannot cause FASD births. No Finnish studies on prevalence of unintended pregnancies continuing to birth was found. However, stable abortion rates in Finland between 1990-2022 suggest a relatively stable rate of unintended pregnancies over time. In Northern Europe 17% of births have been estimated to be results of unintended pregnancies [55]. In a Danish cohort study 23.9% of continuing pregnancies were not classified as very planned or fairly planned [56].

Continued alcohol use before pregnancy recognition (BrP) is not solely attributable to unintended pregnancies. In Australia, more than 60% of women reported alcohol use before recognizing their pregnancy, despite 73% indicating that the pregnancy was planned [57]. Binge drinking was also common prior to pregnancy recognition, and higher socioeconomic status was associated with an increased likelihood of prenatal alcohol exposure (PAE) [57]. Similar patterns have been observed in Denmark, a Nordic country comparable to Finland, where studies indicate that binge drinking before pregnancy recognition occurs even among women with planned pregnancies [56,58].

Pregnancy recognition timing has not been studied in Finland. In the US recognition timing remained stable between 1990-2012 [9], with a mean at 5.5 weeks and 23% recognized at 7 weeks or later [9]. These numbers imply that majority of women who continue alcohol use until awareness of pregnancy are at risk for any PAE during gastrulation.

Very late pregnancy awareness is rare but associated with very high risk for PAE and subsequent FASD in populations with prevalent alcohol use. In Germany 1/500 of pregnancies were recognized after the 20 weeks and 1/2500 only at birth [59]. A similar prevalence of cryptic pregnancies was reported in the UK in the 2020’s [60]. As both biomarker and self-reporting studies on PAE have typically been conducted before gestational week 20, the number of very late awareness must be included separately.

PAE prevalence among pregnancies with late awareness was assumed as high as alcohol use prevalence among women of fertile age despite of being likely higher as AUD increased risk for both unintended pregnancies and late awareness [61].

Based on above-mentioned data, assumption for constants in continuing pregnancies were:

- 20% are recognized after 6+0 (*L*=0.2), at least four weeks after conception
- 0.2% (=1/500) are recognized at 20+0 or later (*L*_x_=0.002)

#### Country multiplier M

To find the country multiplier *M*, we compared each country’s FASD prevalence (*Φ*) estimates to *A*_h_ (WHO GHO) for that country using mean for the years of gestation of the study population. If available, we used the observed prevalence (number of diagnosable FASD children/evaluated participants) in each study. If the actual observed prevalence was not reported, we used reported estimated prevalence. For countries with more than one study, we used the mean of the given estimates.

Due to the differences among the included studies, we applied weights for each study. First, we ranked the studies according to the diagnostic guidelines used. As studies applying Canadian guidelines had considerably wider range and more heterogenous reporting and analysis methods, studies using IOM guidelines, both 2005 version and 2016 version, were prioritized. Those IOM criteria have also been applied in Finland, 2005 guidelines before 2016, and 2016 guidelines since. Thus, we assigned studies using IOM 2005 double the weight of the studies using Canadian guidelines, and IOM 2016 studies double the weight of the IOM 2005 studies. Resulting methodological weights *w*_m_ ranged from 1 to 4.

Next, we assigned weights to the studies according to their size. Size weights *w*_s_ were simply the number of participants in each study divided by the total number of participants in all studies. In none of the studies all the participants completed the whole protocol. We defined the completion weights *w*_c_ simply as the proportion of participants that completed the whole protocol in each study. Total weights of the studies were the product *w*_m_*w*_s_*w*_c_. which were normalized to final weights *w*_i_ by

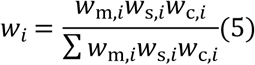

where *Φ_i_* is the observed FASD prevalence for each study. *A_i_* is the country specific binge drinking prevalence corresponding to the studied cohort. In addition to the estimated *M*, we calculated conservative *M_c_* by assuming true values of *Φ*_i_ and *A*_i_ to be 20% lower.

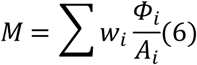

### Estimation of immigrant children with FASD

As population growth in Finland during the last two decades has been driven by immigration, we also conservatively estimated the number of children with FASD who immigrated to Finland between 1990 and 2025. FASD prevalence among immigrants may differ from that of the general population in their countries of origin, but this has not been studied, at least not in the Finnish context.

The largest immigrant groups in Finland during 1990-2020 originated from Estonia and Russia, and since 2022 from Ukraine, all having substantial estimated FASD prevalence (2.8% for Estonia, 2.9% for Russia and 2.7% for Ukraine) according to the global estimate study [14]. Even though those prior FASD estimates appear very conservative according to more recent in-school study observations, we use them to approximate FASD prevalence (*Φ*) in immigrant children from those countries. Conservative estimates here are plausible, assuming that AUD prevalence among mothers who migrate is probably lower compared with those who remain. Other major countries of origin (e.g. Iraq, India, China, Somalia, Syria and Afghanistan) had low estimated FASD prevalences and were here assumed as negligible to further ensure a very conservative overall estimate among all immigrant children. Children were included in the Finnish FASD population in the year of immigration.

A distinct immigration category is international adoption, which peaked in Finland during the early 2000s. It has been shown that at least 50% of children adopted from Eastern Europe have FASD [62,63]. Given the exceptionally high FASD prevalence reported in disadvantaged populations in South Africa [14,64], similar or even higher rates are plausible among adoptees from South Africa, a major country of origin for children adopted to Finland. Furthermore, maternal risk factors associated with PAE, including unplanned pregnancy, psychosocial stress and socioeconomic adversity, are likely overrepresented in populations relinquishing children for international adoption. Consequently, even in countries with low general FASD prevalence, rates among adoptees may be substantially higher.

We therefore assumed FASD prevalence of 50% among adoptees from Eastern Europe and South Africa, and 20% among adoptees from Colombia, Philippines and Thailand, where substantial alcohol use among women in high-risk populations have been reported [65–68].

Data on alcohol use among women in those countries during the period of active international adoption to Finland (1995-2010) is limited. To maintain a conservative approach, we assumed negligible FASD prevalence among adoptees from other countries of origin with substantial number of adoptees (China, India and Ethiopia), even though it has been stated that FASD should be considered in any adopted child having developmental challenges [69]. Numbers of immigrant children in Finland included in FASD estimations for each 5-year period are reported in Table 4.

**Table 4.** Children immigrated to Finland from major countries of origin with considerable FASD risk, estimated number of children with FASD.

| Country | 1990-1994 | 1995-1999 | 2000-2004 | 2005-2009 | 2010-2014 | 2015-2019 | 2020-2024 |
| --- | --- | --- | --- | --- | --- | --- | --- |
| Immigrated with parents |  |  |  |  |  |  |  |
| Russia (R) | 1546 | 2684 | 2445 | 2309 | 2584 | 1818 | 3157 |
| Estonia (E) | 2126 | 1057 | 991 | 2518 | 5383 | 2142 | 1388 |
| Ukraine (U) | 56 | 124 | 134 | 140 | 202 | 442 | 8925 |
| Estimated $I_i(t)$ *<br>=$0.029R+0.028E+0.027U$ | 106 | 111 | 103 | 140 | 231 | 125 | 521 |
| International adoptees |  |  |  |  |  |  |  |
| Eastern Europe (EE) | 198 | 443 | 312 | 235 | 151 | 42 | 41 |
| South Africa (SA) | 1 | 1 | 67 | 150 | 179 | 109 | 80 |
| Colombia (C) | 97 | 118 | 173 | 78 | 45 | 38 | 28 |
| Thailand (T) | 43 | 118 | 187 | 197 | 103 | 82 | 118 |
| Philippines (PH) | 0 | 4 | 26 | 82 | 66 | 44 | 27 |
| Estimated $I_a(t)$ *<br>=$1/2(EE + SA)+$<br>$1/5(C+T+PH)$ | 128 | 270 | 267 | 264 | 208 | 109 | 96 |
| *Rounded to complete children |  |  |  |  |  |  |  |

Following this, we calculated FASD prevalence estimates among immigrant children as

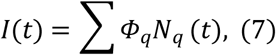

where q represents the country. *Φ*_q_ and *N*_q_ are the country specific FASD prevalences and yearly immigration numbers, respectively.

### Used software

We implemented the models and produced the figures with R, version 4.5.2 [70].

## Results

According to available research data, alcohol use among women in Finland remained high and stable with tendency to binge drinking and a slight decrease since 2008 (Figure 1). Regardless, prevalence of binge-drinking among women was constantly higher in Finland compared to the countries in the FASD prevalence studies.

**Figure 1.**
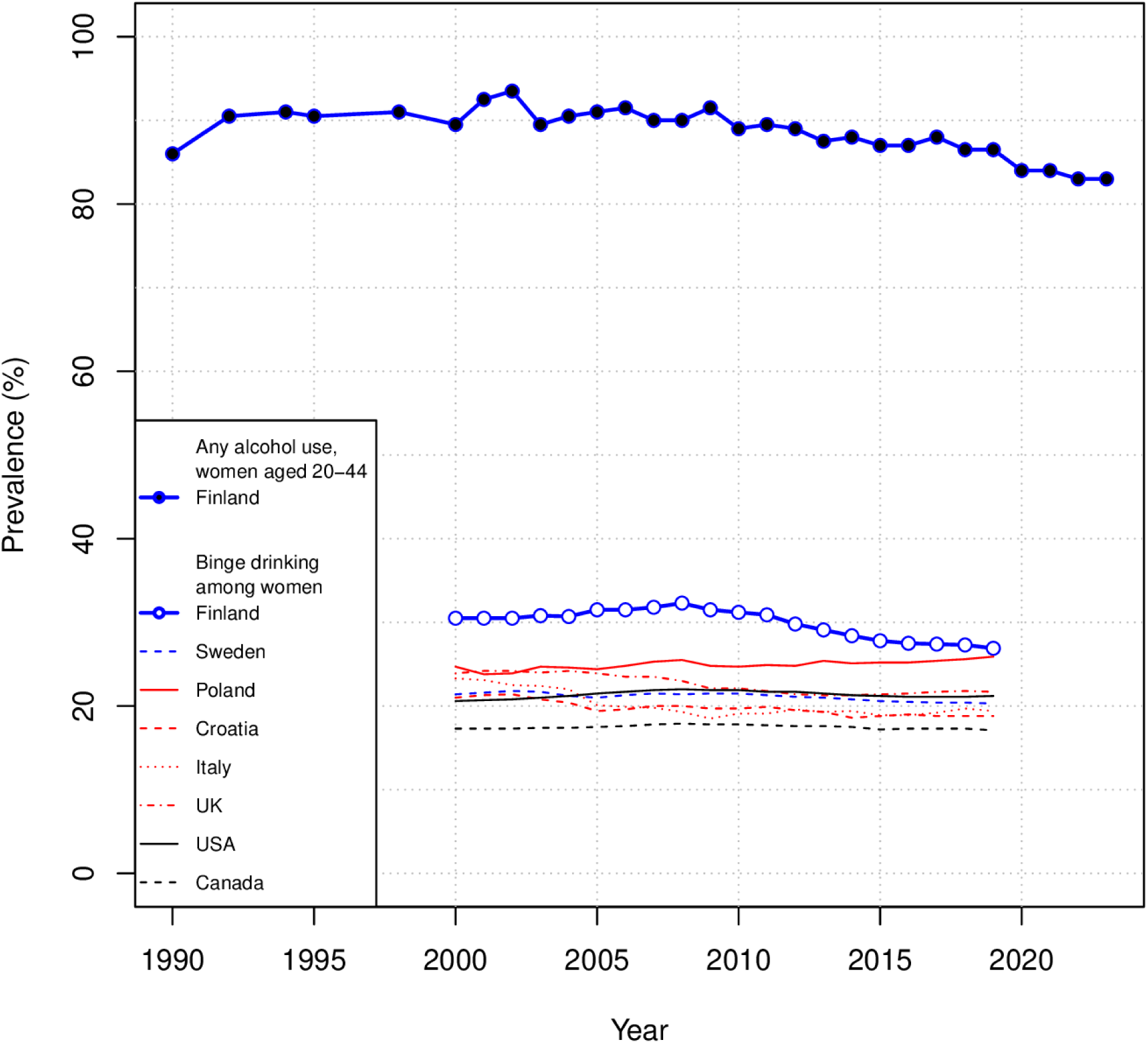
Alcohol use by women of child-bearing age in Finland and the binge drinking prevalence among women in Finland and seven countries of the FASD prevalence studies. Heavy episodic alcohol use within last 30 days data (WHO)[54] includes all women. Any alcohol use data (Statistics Finland) shows the prevalence of any alcohol use of women aged between 20-44 years in Finland. The colors group countries as Nordic, other European, or North American.

Self-reported rate of any PAE has decreased significantly since official recommendation for total abstinence during pregnancy 2004 (Figure 2A), contradicting general statistics on alcohol use that present a more stable consumption. Accordingly, the heavy alcohol consumption parameter *α* had only a moderately decreasing trend (Figure 2B).

**Figure 2.**
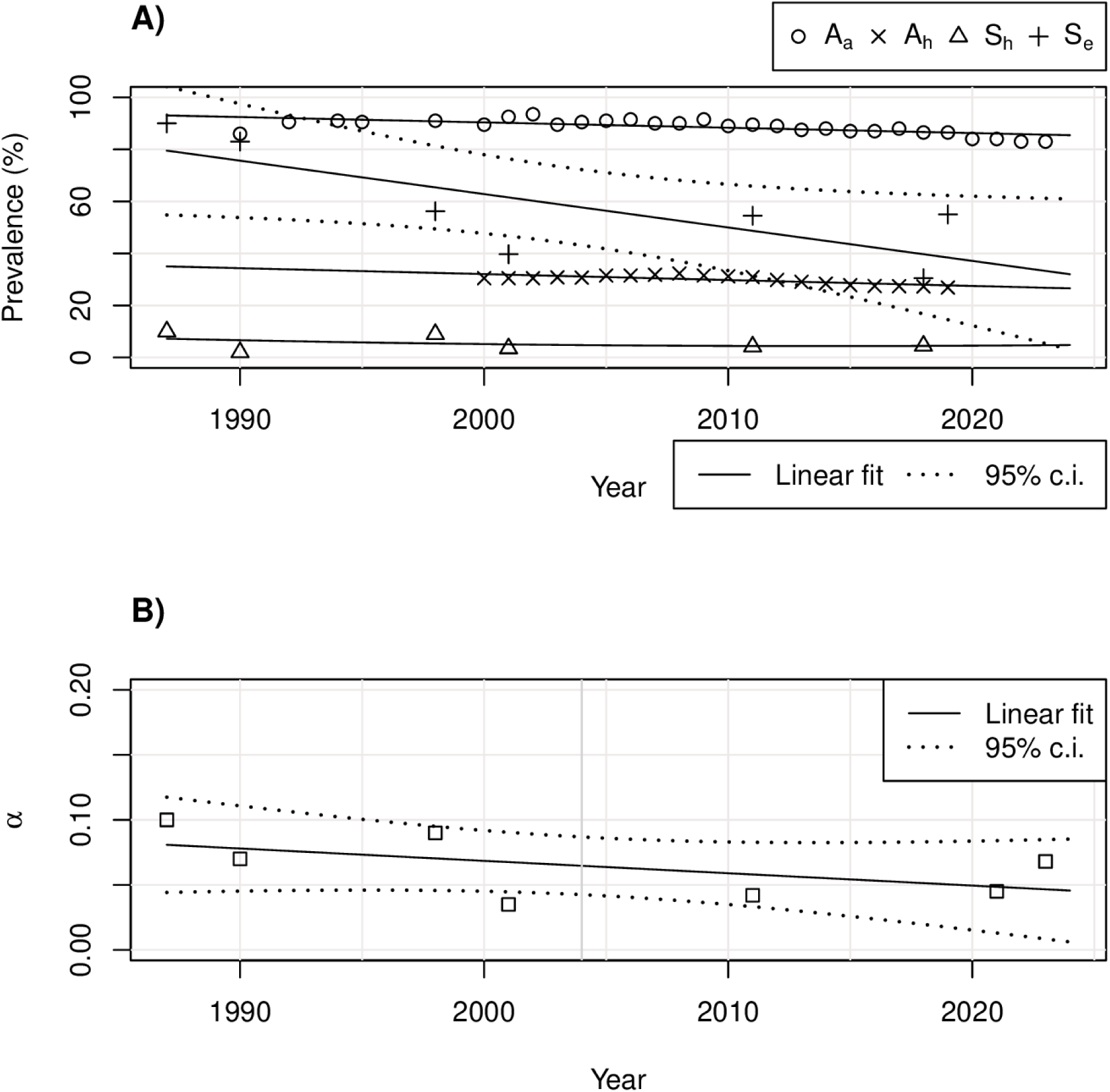
Alcohol consumption trends and the heavy alcohol consumption parameter α. A) Self-reported alcohol use during pregnancy (S_e_) shows substantially different trend compared with other alcohol use indicators, such as general alcohol use prevalence by women aged 20 – 44 (A_a_), binge drinking (A_h_) and self-reported heavy alcohol use during pregnancy (S_h_). B) Linear approximation of the heavy alcohol use during pregnancy parameter α decreases with time, though the biomarker studies after year 2020 suggest either more gradual decrease or even a possible turn in the trend. Squares show actual alcohol use prevalence data points.

The proportion of children with any self-reported PAE, including before recognition of pregnancy has declined from 75% (uncertainty range 60%-85%) in the 1990s to 32% (26%-38%) in early 2020s. (Figure 3). Incidence of estimated heavy PAE during pregnancy has declined from 9% (7%-11%) in the 1990s to 6% (4%-8%) in the early 2020s (Figure 4).

**Figure 3.**
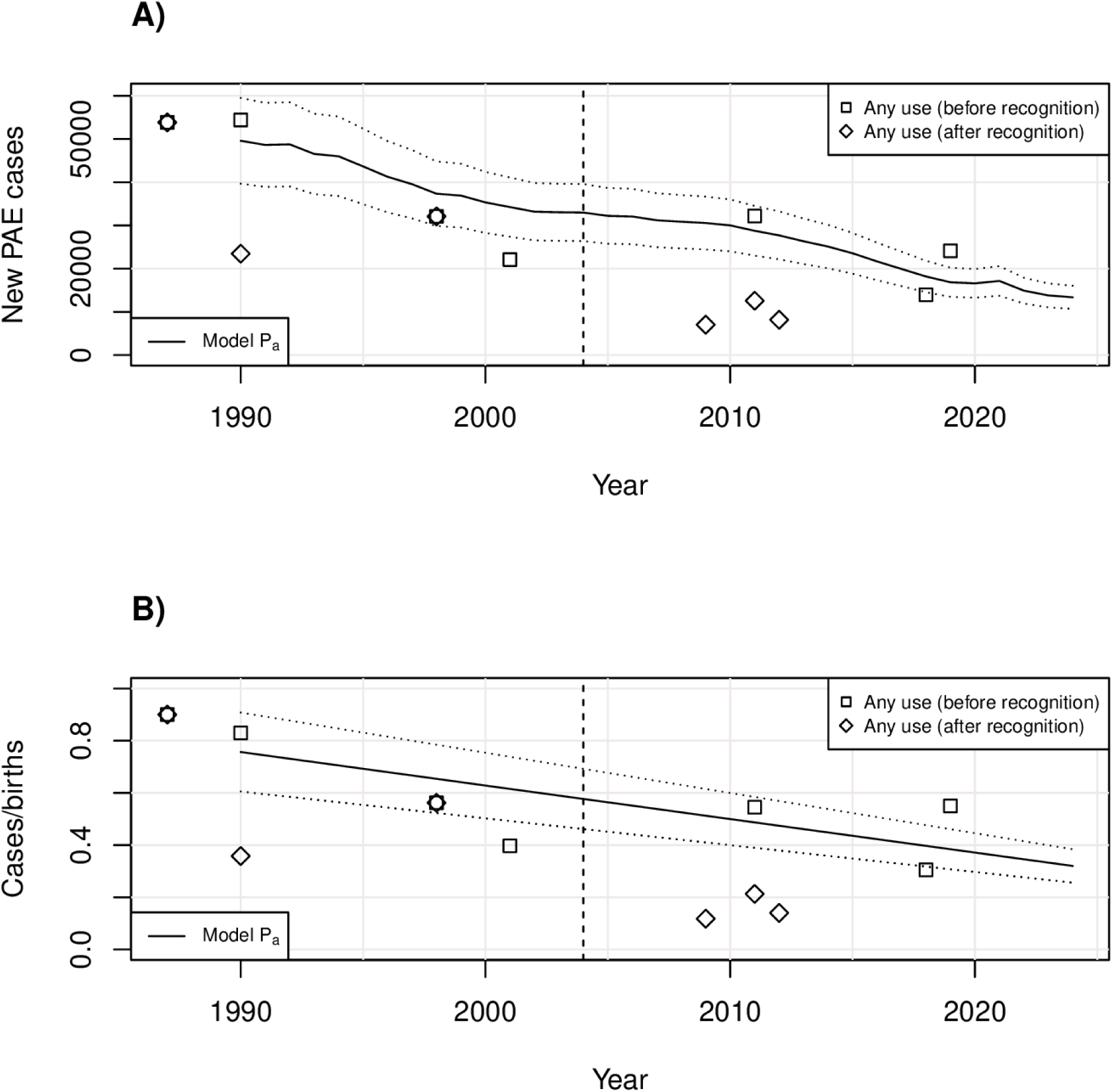
Prevalence of self-reporting any alcohol use by pregnant women and estimated associated any PAE incidence. A) Solid line marks the mean estimated yearly increase in Finnish children with any PAE effects. Dotted lines frame ±20% uncertainty range associated with the self-reported data. Points refer to specific data points according to the studies, categorized to usage before and after the recognition of pregnancy. B) The same estimation and data points as percentages of each annual birth cohort. Dashed vertical line marks the year 2004, when official recommendation of total abstinence was made.

**Figure 4.**
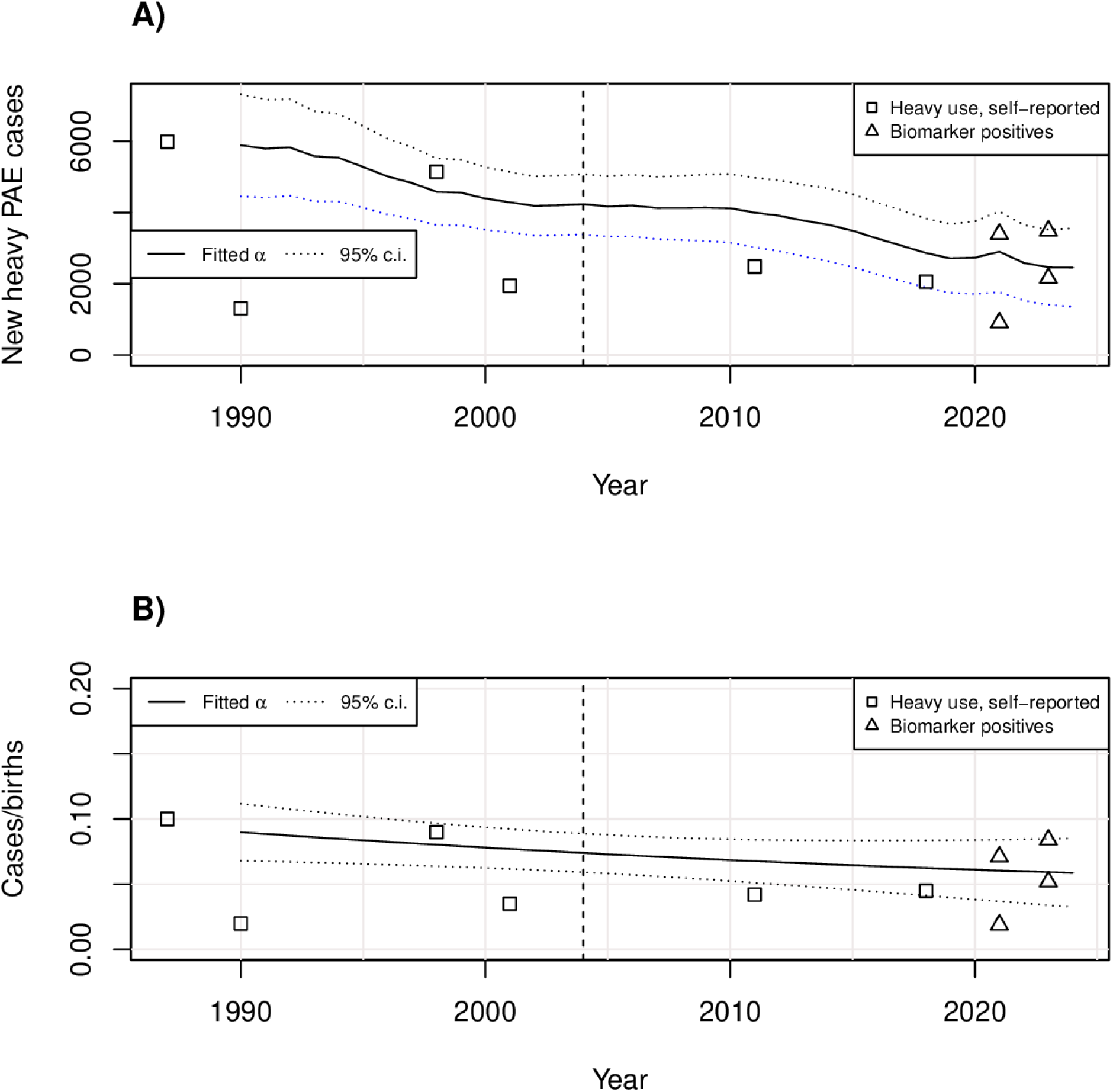
Prevalence of heavy alcohol use by pregnant women and estimated associated heavy PAE incidence. A) Solid line marks the mean estimated yearly increase in Finnish children with severe effects of PAE. Dotted lines frame ±20% uncertainty range associated with the self-reported data. Points refer to specific data points according to the studies, categorized to self-reported data and biomarker studies including both cut-offs when two have been reported. B) The same estimation and data points as percentages of each yearly birth cohort. Dashed vertical line marks the year 2004, when official recommendation of total abstinence was made.

Internationally, the prevalence of heavy episodic drinking in the past 30 days among women (A_h_) using mean for each country was correlated (Pearson 0.70, with p=0.079 due to low number of countries with studies) with FASD prevalences in active case ascertainment studies (Figure 5A). Estimated country multiplier M that links FASD prevalence to the prevalence of heavy episodic drinking in a country was approximated at 0.298. Using that simple internationally derived M indicates remarkably high incidence of FASD in children born in Finland between 1990 and 2025 fluctuating from 8.2 to 10.2% (Figure 5B) with peaks in the early 1990s, 2008-2010 and a smaller increase in 2020-2021 during otherwise declining trend since 2011. Using a more conservative estimate with 20% reduced M of 0.198 indicates a FASD incidence fluctuating from 6.8% in early 1990s to 5.6% in 2020s with decreasing trend since 2011, with the exception of the Covid pandemia period (Figure 5B). Model L based on prior estimated ratio of any PAE/13 indicates FASD prevalence with steeper decrease from 6% to 3% depending on year (Figure 5B). As model L depends on self-reported PAE, it deviates from model M estimates since the introduction of recommendation of abstinence during pregnancy.

**Figure 5.**
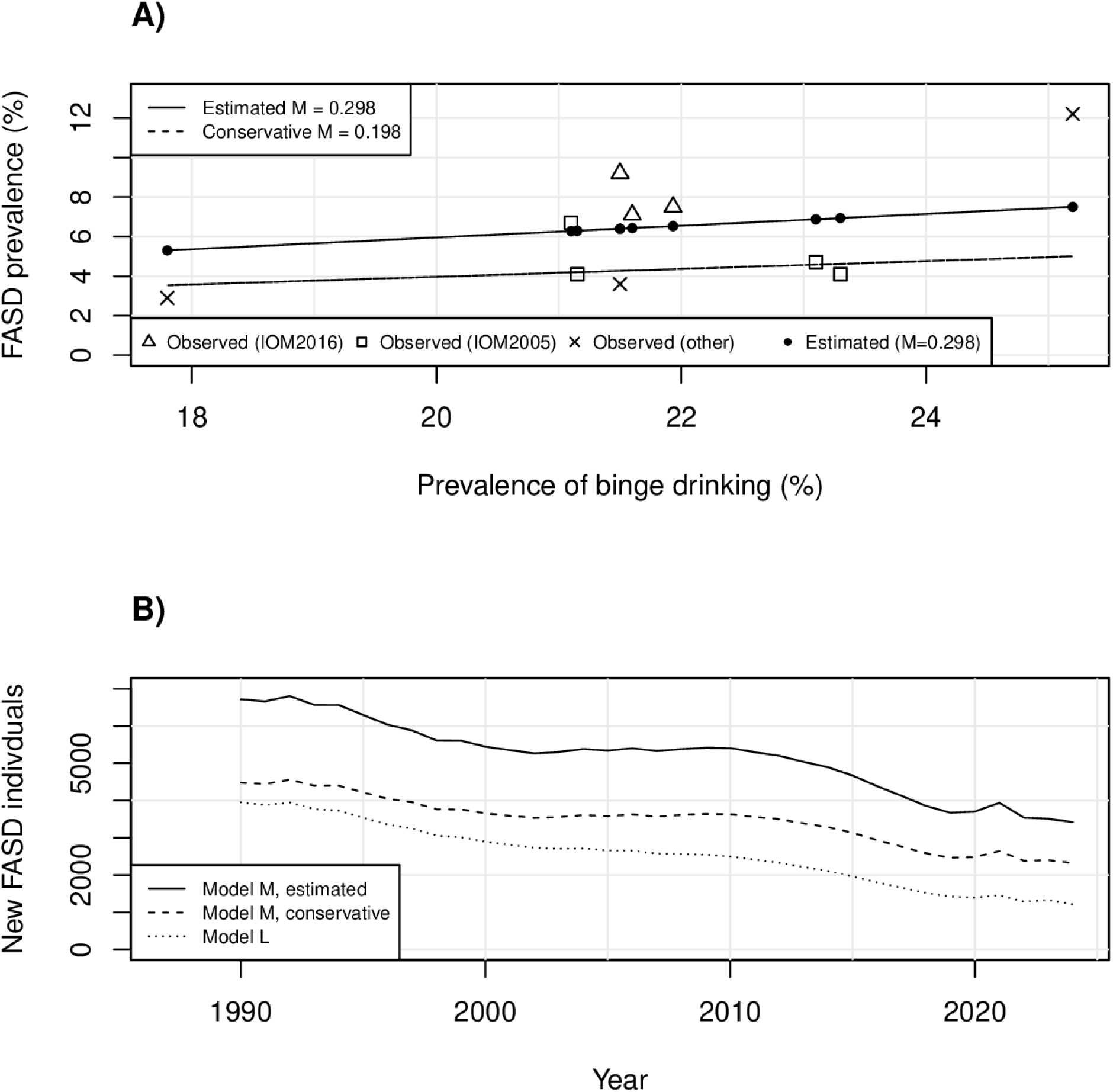
Estimation of new FASD cases in Finland 1990-2025. A) Generation of multiplier M using observed FASD prevalences and prevalences of binge-drinking among women Points refer to FASD prevalences in international studies and the corresponding estimates given by Model L without immigration data. B) Estimated yearly increase in children eligible for FASD diagnosis in Finland. Model L follows closely conservative M estimate until the recommendation of abstinence 2004.

The decreasing number of continuing alcohol use after recognition of pregnancy and slight reduction in alcohol use among women in fertile age in general have contributed to decreasing estimated numbers of children born with FASD during the 2020’s.

The annual number of new children with FASD in Finland has declined also due to the increasing proportion of life-time abstainers due to religious or other reasons among those giving birth, declining birth rate and decreasing number of international adoptees. Still, approximately 1500- 2500 new children with FASD are estimated annually using conservative numbers.

Immigrant children with FASD have comprised a small minority of approximately 2% (1-4% depending on year) of new cases with FASD. An estimated 2679 children with FASD immigrated to Finland in 1990-2024. Approximately half of the immigrated children with FASD were internationally adopted (1342) and half (1337) immigrated with their parents. Both estimates are conservative.

## Discussion

The estimated incidence of any PAE, including exposure prior to pregnancy recognition (32-75%, range 26-85%), heavy PAE (6-9%, range 4-11%) and subsequent FASD (5-7%, range 3-10%, depending on the model) were remarkably high for Finland. These estimates are substantially higher than the only previous attempted estimate reporting a FASD prevalence of 1.2% [14]. That estimate was largely based on assumption that Finland’s relatively high socioeconomic status (SES) would be associated with low FASD prevalence, despite evidence of high overall alcohol use and particularly high rates of binge drinking among women. Moreover, there is lack of evidence that a higher SES reduces PAE; rather, some studies suggest it may actually be associated with a higher likelihood of alcohol use during pregnancy [33,57,58]. It should also be noted that the earlier estimate was conducted before the publication of more recent in-school active case ascertainment studies and modern biomarker findings, both of which have revealed substantially higher prevalence estimates than earlier methodological approaches.

The decreasing trend observed in both PAE and FASD in the present study is encouraging. However, this trend may partly reflect methodological artefacts. European biomarker studies have demonstrated that self-reported alcohol consumption during pregnancy is very frequently underreported [16,18]. Although true alcohol abstinence during pregnancy has likely increased, strict abstinence recommendations contribute to underreporting, thereby complicating the identification of PAE.

Patterns of alcohol consumption among women are relevant when considering risks for PAE and FASD. Heavy episodic drinking increases risk of inadvertent heavy PAE during early pregnancy, before pregnancy recognition. This has been demonstrated also in a Danish cohort study [56]. Thus, binge drinking increases especially the risk of early pregnancy birth defects. This type of alcohol use pattern is also associated with a higher risk of alcohol use disorder. Both of these compounding risks contribute to an elevated FASD risk in populations with this traditional drinking pattern. When compared with countries where in-school FASD prevalence studies have been conducted, Finland has had a higher rate of binge drinking among women, likely resulting in a higher FASD risk including alcohol-related birth defects. This common drinking pattern may explain the remarkably high prevalence of congenital malformations in the control population of our previous study [61].

Our estimates are plausible and purposely conservative compared with results of European in-school active case ascertainment studies. It should be noted that women’s alcohol use pattern in Finland during much of the study period most resembled those in Poland, where the observed FASD prevalence rate was 12.2% among participants of the in-school study [33]. Sweden represents the culturally closest country to Finland among those where FASD prevalence studies have been conducted, and the Swedish study reported a conservative FASD estimate of 5.5% and an observed prevalence among 9.2% of the participants. During that study period, both countries had well-developed antenatal care systems and similar abstinence recommendations during pregnancy. However, women’s heavy episodic alcohol use prevalence in Sweden was significantly lower than that of Finland. This is consistent with our slightly higher estimation for the FASD prevalence in Finland during the corresponding period compared to the conservative Swedish estimate [35].

Although not all children with FASD are born to mothers with AUD, many are. A comparison of our FASD estimates with indicators of maternal AUD during pregnancy supports this relationship. In Finland, all pregnant women complete the Alcohol Use Disorder Identification Test (AUDIT) at their first prenatal visit (gestational week 8-11). Women with high AUDIT scores are referred to specialized HAL clinics (Huumeet, Lääkkeet, Alkoholi [Drugs, Medications, Alcohol]). In 2017, a total of 1094 women attended HAL clinics [71], corresponding to approximately 2.2% of all pregnancies. It has been estimated that only about one third of women with substance use disorder are identified, referred to and subsequently attend HAL clinics [72]. This would imply that approximately 6.6% of pregnant women may have problematic substance use, aligning with the results from Pajulo et al. [73], which showed that 6.4% of pregnant women had substance dependency. Although not all patients with substance use disorder primarily use alcohol, this estimate is broadly consistent with our modeled FASD incidence for that year.

The low, but notable estimated number of immigrant children with FASD is consistent with both clinical observations and previous research. A Finnish study of diagnosed FASD cases reported a higher rate of immigrant children, both international adoptees and children immigrated with their parents, among FASD diagnosed [74]. Furthermore, research on adopted children in Finland has shown that one third of all internationally adopted children, and one half of those adopted from Eastern Europe, experience learning difficulties [75], which is consistent with the assumptions used in our estimates.

## Limitations

The primary limitation of this study is the lack of comprehensive objective and specific data. Only a small number of fully anonymous or biomarker-based studies on PAE have been conducted in Finland, resulting in fragmented data and substantial degree of uncertainty in estimates. The stigma associated with alcohol use during pregnancy likely contributes to systemic underreporting in self-reported data. However, because over-reporting is unlikely, self-reported alcohol use can reasonably be interpreted as representing minimum prevalence levels, leading to our estimates being conservative.

Biomarkers of alcohol exposure also tend to provide conservative estimates. Although biomarkers offer more objective evidence than self-reporting, most have short exposure detection windows. Only meconium or maternal hair samples could detect alcohol exposure during longer periods, both retrospectively. Furthermore, many currently used biomarkers were developed to detect levels of alcohol use associated with health risks to the drinker, which limits their sensitivity for detecting lower levels of exposure, that could nevertheless be harmful to the more vulnerable developing embryo or fetus.

The results of the present study also depend on several assumptions derived from the best available evidence. For example, as there are no recent data on prevalence of unintended pregnancies in Finland, our assumption is based on older and international numbers. However, unplanned pregnancies have been common among women with heavy alcohol use [61].

The heterogeneity of the underlying studies used in the modeling process introduces additional uncertainty; a limitation encountered in all review-based approaches.

Finally, regional variations in FASD prevalence may be substantial within countries, as demonstrated in studies conducted in the United States. This variation introduces potential error when comparing national binge-drinking rates among women with prevalence estimates derived from more geographically limited FASD studies. For this reason, prevalence estimates based on multiple studies within a country are likely to provide more reliable comparisons.

## Conclusion

PAE and FASD have likely represented, and continue to represent, significant public health concerns in Finland. However, several demographic and behavioral trends, including declining birth rates, decreasing alcohol consumption, increasing awareness of alcohol-related harms and growth of immigrant populations with lower alcohol consumption, may contribute to a gradual reduction in the burden of PAE-related harm among children. Nevertheless, even if complete abstinence during pregnancy were achieved in the future, a substantial adult population affected by PAE and undiagnosed FASD would remain.

To improve the accuracy of current prevalence estimates in Finland, further studies are needed. For PAE prevalence, population-based studies using meconium or maternal hair EtG biomarkers would provide more objective estimates. For FASD prevalence, in-school active case ascertainment studies would be particularly valuable.

## Data Availability

All relevant data are within the manuscript and its Supporting Information files.

## Acknowledgements

We thank Mr. Trevor Yoak for his excellent help at the preparation of the manuscript.

## Supporting Information

S1 Dataset. Parameters 1987 – 2024. Zipped datasets (4 csv files) to reproduce the results.

S2 Source. R source file. R script to reproduce the results.

